# A Phenomenological Analysis of COVID-19

**DOI:** 10.1101/2021.05.23.21249471

**Authors:** A. O. Macchiavelli, G. V. Martí, J. G. Martí

**Affiliations:** Nuclear Science Division, Lawrence Berkeley National Laboratory, Berkeley, CA 94720, USA; Laboratorio Tandar, Comisión Nacional de Energía Atómica, Av. Gral. Paz 1499, BKNA 1650, San Martín, Argentina; Escuela de Ciencia y Tecnología, Universidad Nacional de San Martín, Argentina

**Keywords:** COVID-19, Mathematical modeling, Compartimental models, Pandemic, Governmental actions, Epidemic, Outbreak, SIR-SIS models and SEICRD extended model

## Abstract

A phenomenological analysis of the time evolution of some COVID-19 data in terms of a Fermi-Dirac function is presented. In spite of its simplicity, the approach appears to describe the data well and allows to correlate the information in a universal plot in terms of non-dimensional or reduced variables *N*_*r*_ = *N* (*t*)*/N*_*max*_, and *t*_*r*_ = *t/*Δ*T*, with *N* (*t*) being the total number of cases as a function of time, *N*_*max*_ the number of total infected cases, and Δ*T* the diffuseness of the Fermi/Dirac function associated with the rate of infection. The analysis of the reported data for the first outbreak in some selected countries and the results are presented and discussed. The approach is also applicable to subsequent waves. Support of our framework is provided by the SIS limit of the SIR model, and simulations carried out with the SEICRD extension.

## 1. INTRODUCTION

The worldwide outbreak of COVID-19 has triggered a large number of studies. In fact, a search in medRxiv, bioRxiv [1], and arXiv[2] (across all fields) returns more than 8800, 2400 and 2900 articles respectively, that have been posted since the beginning of the year 2020 ^1^. It goes without saying that this impressive number of manuscripts clearly reflects the scientific urgency to understand the behavior of this novel virus and to develop and to assess several models that can shed light on its outbreak and make reliable predictions of its dynamical evolution.

In this work, as a contribution on the topic of mathematical modeling, we would like to point out that the available data [3, 4] for some selected countries could be described by a Fermi/Dirac (F/D) function, introduced in physics by Fermi and Dirac to describe the distribution of particles over energy states in systems consisting of many identical particles that obey the Pauli exclusion principle [5–7]. This type of function is also extensively used in nuclear physics to parametrize the nuclear potential (Wood-Saxon potential) in studies of structure and reactions [8].

Our phenomenological analysis of the first outbreak of the pandemic, presented in Sections II and III, provides a reasonable description of the data and the evolution of the pandemic without relying on any specific epidemiological model. An interesting finding is that the data follows a universal behavior based on dimensionless variables that allows, in a simple manner, to relate the response of the pandemic to measures adopted to counteract the spread of the virus. While we mainly focus our discussions on the first outbreak, the method can obviously be extended for the subsequent ones as briefly presented in Section IV.

## II. THE PHENOMENOLOGICAL APPROACH

We make the conjecture that the data can be described by an analytical function of the F/D type:

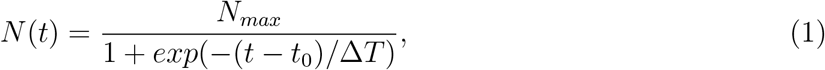

where *N* (*t*) is the cumulative number of infected cases, *N*_*max*_ is the total number of cases, Δ*T* is the diffuseness of the F/D function and *t*_0_ is a reference time when 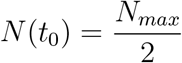 The use of this type of function can be justified by considering the first derivative of Eq. 1:

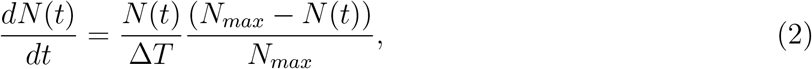

showing that the rate of infections at a given time is proportional to the product between the infected and non-infected cases. In Ref. [9], the authors considered this type of function in an analysis of the early pandemic in Sweden. Furthermore, the so-called SIR (Susceptible, Infectious, Recovered) model [10], widely used in epidemiology to study virus transmissions, consists of a system of three time-dependent variables: *I*(*t*), the number of total infected individuals at a given time, *S*(*t*) the number of individuals susceptible of contracting the infection, and *R*(*t*) the cumulative number of recovered individuals. In a closed system with a constant population size, *N*_*max*_, these variables satisfy the following equations:

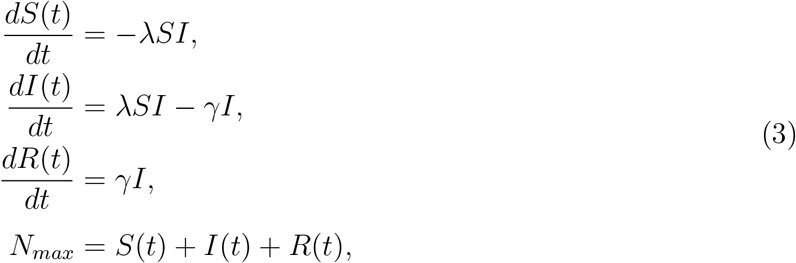

where *λ* is the transmission rate and *γ* the recovery rate. If the recovery rate is very low during the pandemic time interval (*γ* ≈ 0), we can approximate the infected cases by:

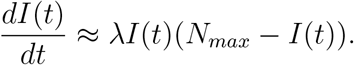

This limit is known as the SIS (Susceptible - Infected - Susceptible) model with the solution above readily recognized in Eq. 2, with *λN*_*max*_ = 1*/*Δ*T*.

In the SIR model the basic reproduction number, ℛ_0_, is given at the outset of the epidemic by: *λN*_*max*_*/γ* = 1*/*(*γ*Δ*T*) (See the nice discussion in Ref. [11]). If for a given population ℛ_0_ *>* 1 the infection will spread exponentially, while if ℛ_0_ *<* 1 the infection will progress slowly and eventually die out. The value of ℛ_0_ for COVID-19 is estimated to be between 2 and 4. An effective reproduction number, ℛ, is often used to reflect the state of the epidemic at any given time. ℛ = ℛ_0_(1-*P*_*i*_) where *P*_*i*_ is the proportion of the population who is immune at that time [12].

## III. ANALYSIS

As mentioned above, we focus our analysis on the first outbreak and start our discussion with data^2^ from China^3^, shown in Fig. 1. A fit of Eq. 1 reproduces well the empirical distributions for the cumulative and daily number cases given by Eq. 2. A similar procedure was applied to the data from a number of selected countries listed in Table I, including the city of New York in the USA, which will be further discussed in Section IV. The fit parameters are also given in Table I.

**FIG. 1:**
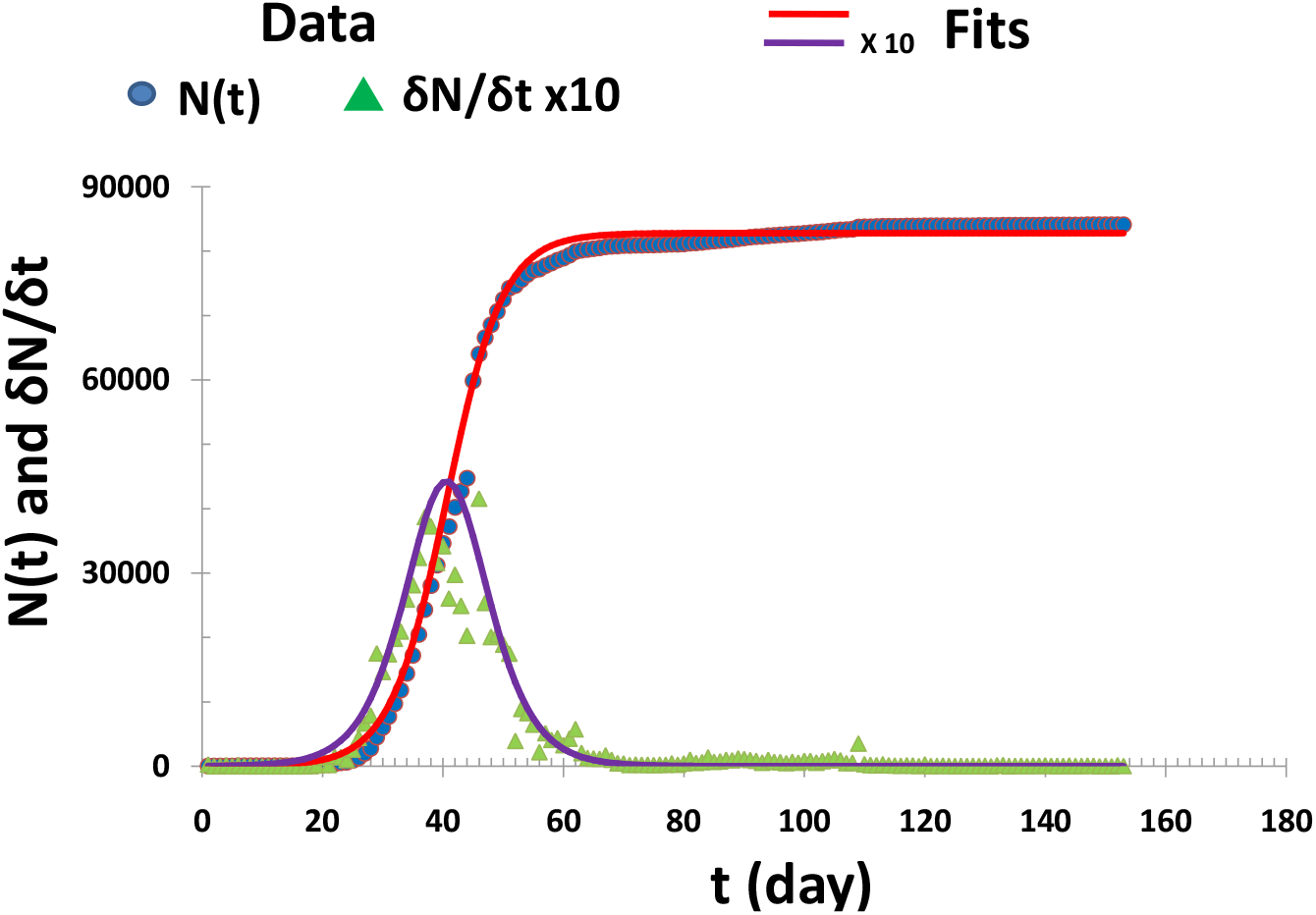
Number of cumulative cases *N* (*t*) as reported in China compared to the fit of an F/D function, with the following parameters, *N*_*max*_ = 82791, *t*_0_ = 40.6 days and Δ*T* = 4.68 days. *dN/dt* and the daily infected cases *δN* (*t*)*/δt* = (*N* (*t* + 1) - *N* (*t*))*/*(1day) are also included, multiplied by a factor of 10.

**TABLE I:** The data were taken from [3] based on the reported data for each selected country since Dec-31, 2019 for China, and since Feb-20, 2020 for the rest of the countries. The data set of New York City were taken from [4]. The cut-off date is May-31, except for Brazil and Perú that the cut-off dates are Sep-30 and Jul-20 respectively.

| Country | $N_{max}$<br>(cases) | $t_0$<br>(days) | $\Delta T$<br>(days) | $PR(t_0)$<br>(cases/day) | $t_{90}$<br>(days) | $N_{90}$<br>(cases) | $t_0(\text{emp})$<br>(days) | $PR(\text{emp})$<br>(cases/day) | $N_{90}(\text{emp})$<br>(cases) |
| --- | --- | --- | --- | --- | --- | --- | --- | --- | --- |
| Brazil | 5156471 | 161.2 | 27.38 | 47082 | 221 | 4640824 | 161 | 69074 | 4732309 |
| Canada | 89693 | 65.9 | 11.74 | 1910 | 92 | 80724 | 74 | 2760 | 81313 |
| China | 82791 | 40.6 | 4.68 | 4423 | 51 | 74512 | 44 | 15141 | 74652 |
| Germany | 173158 | 45.6 | 8.29 | 5222 | 64 | 155842 | 37 | 6294 | 150383 |
| Perú | 314875 | 101.4 | 14.72 | 5348 | 134 | 283388 | 104 | 8875 | 295599 |
| Spain | 228423 | 42.7 | 7.91 | 7219 | 60 | 205581 | 36 | 9181 | 195470 |
| Sweden | 38787 | 69.0 | 14.34 | 676 | 101 | 34908 | 71 | 790 | 35023 |
| United Kingdom | 274336 | 64.3 | 11.37 | 6032 | 89 | 246902 | 52 | 8719 | 246407 |
| USA | 1753980 | 64.8 | 12.38 | 35420 | 92 | 1578582 | 66 | 48529 | 1577287 |
| (New York) | 194746 | 47.4 | 9.14 | 5327 | 68 | 175271 | 46 | 6365 | 170236 |

The values of the diffuseness Δ*T* increase from the lowest value for China, going through the values of the European countries, up to the highest values of the North and South American countries. Our interpretation is that the infection rate is progressively delayed further, as the pandemic progressed and the authorities implemented measures to confront and control it.

Other parameters characterizing the time evolution are worth discussing. One follows from the first derivative of *N* (*t*) that has a maximum at its inflection point: 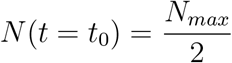 which substituted in Eq. 2, gives the peak rate, 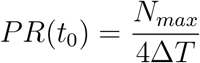. Note that from the numbers in Table I and the relation 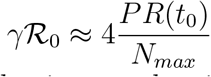, we can estimate a value for *γ* ∼ 0.03*/day*. Measures of the early (asymptotic) behavior are the time, *t*_10_ (*t*_90_), when the number of cases reach 10% (90%) of the maximum, *N*_10_ (*N*_90_). It is simple to show that *t*_10_(*t*_90_) ≈ *t*_0_ - (+)2.2Δ*T*. The parameter *t*_90_ is also given in Table 1 and compared to that obtained from the empirical curves shown in the last three columns (emp). Deviations between the fits and the empirical data vary in the range of 1 to 20% for *t*_0_ and *t*_90_, and from 1 to 5% for *N*_90_. The deviations on *PR*(*t*_0_) are larger, 16-71%, likely due to systematic effects in the reporting of daily cases.

It is more instructive to consider a linearized form of Eq. 1 and plot

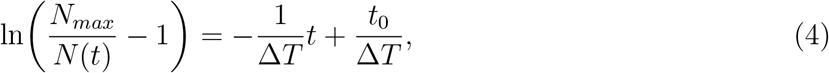

from which we can easily extract Δ*T* from the slope and *t*_0_ from the intercept. The data are presented in this form in Fig. 2 and Fig. 3 for the countries in Table I. The F/D function suggests a universal behavior in terms of dimensionless (or reduced) variables. Introducing:

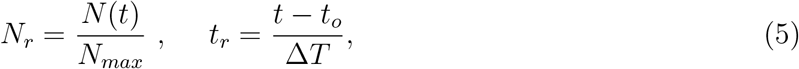

all the data should lie in a common, universal curve. Remarkably this appears to be the case, as seen in Fig. 4. In addition, in terms of these reduced variables Eq. 4 can be written as:

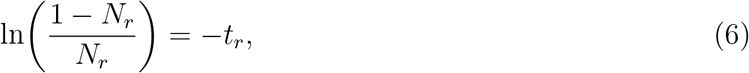

thus, here, the universal curve is a straight line with slope -1 and 0 intercept. The data are presented in this linearized form in Fig. 5.

**FIG. 2:**
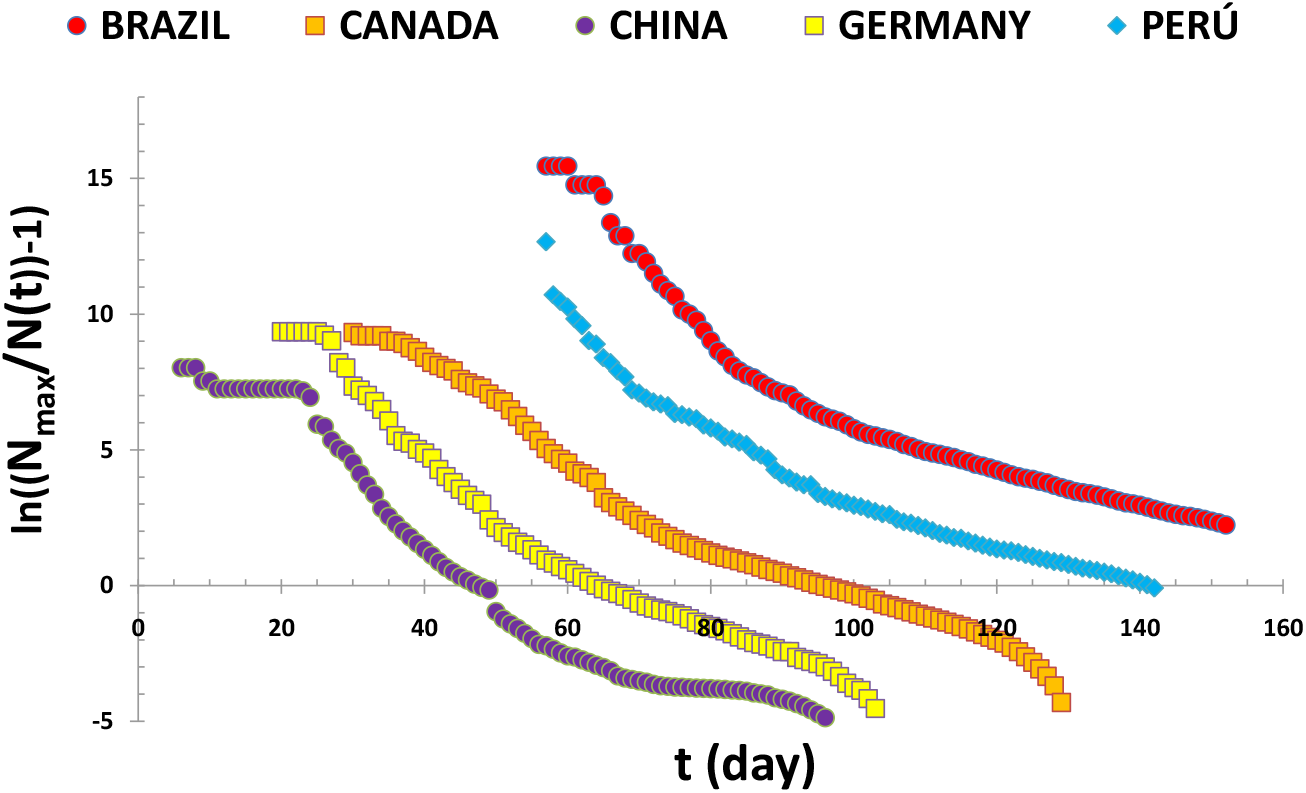
COVID-19 data for China, Germany and some American countries are shown following from Eq. 4. For visual purposes, an arbitrary constant value was added to the times of each data set to avoid overlap of the different straight lines.

**FIG. 3:**
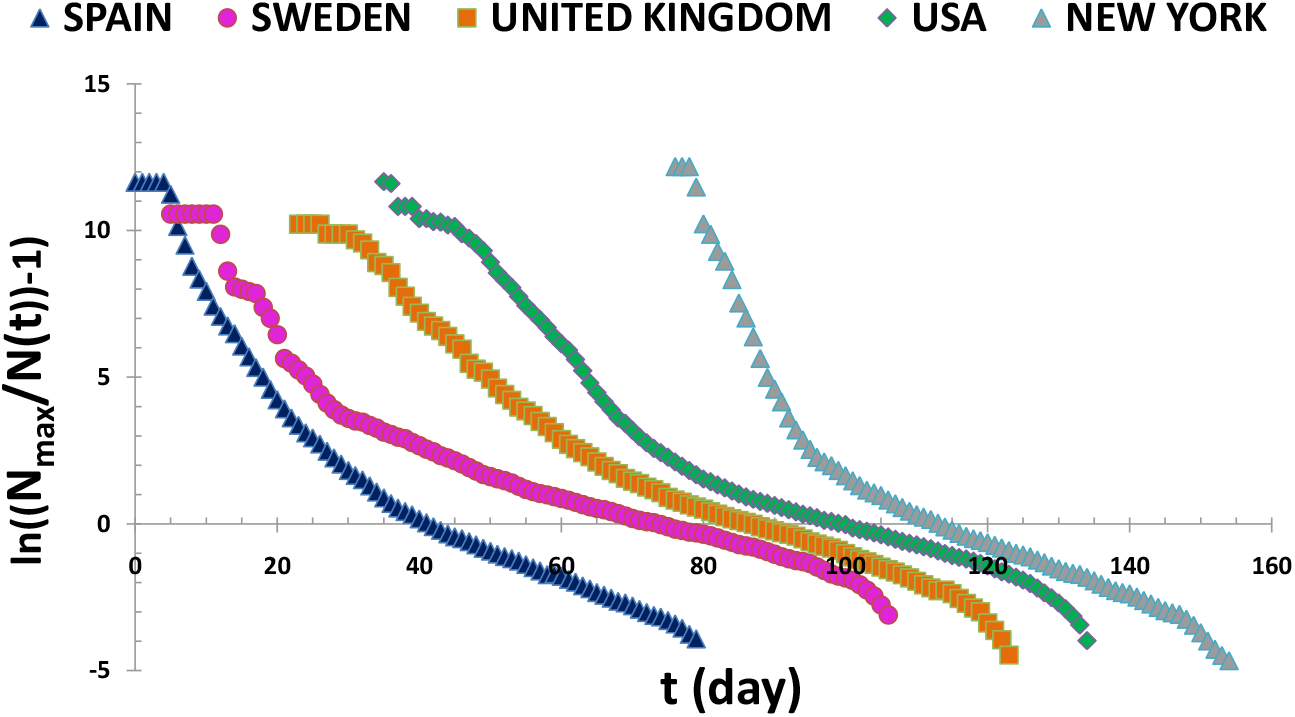
Same as Fig. 2, but for selected European countries, USA and New York City.

**FIG. 4:**
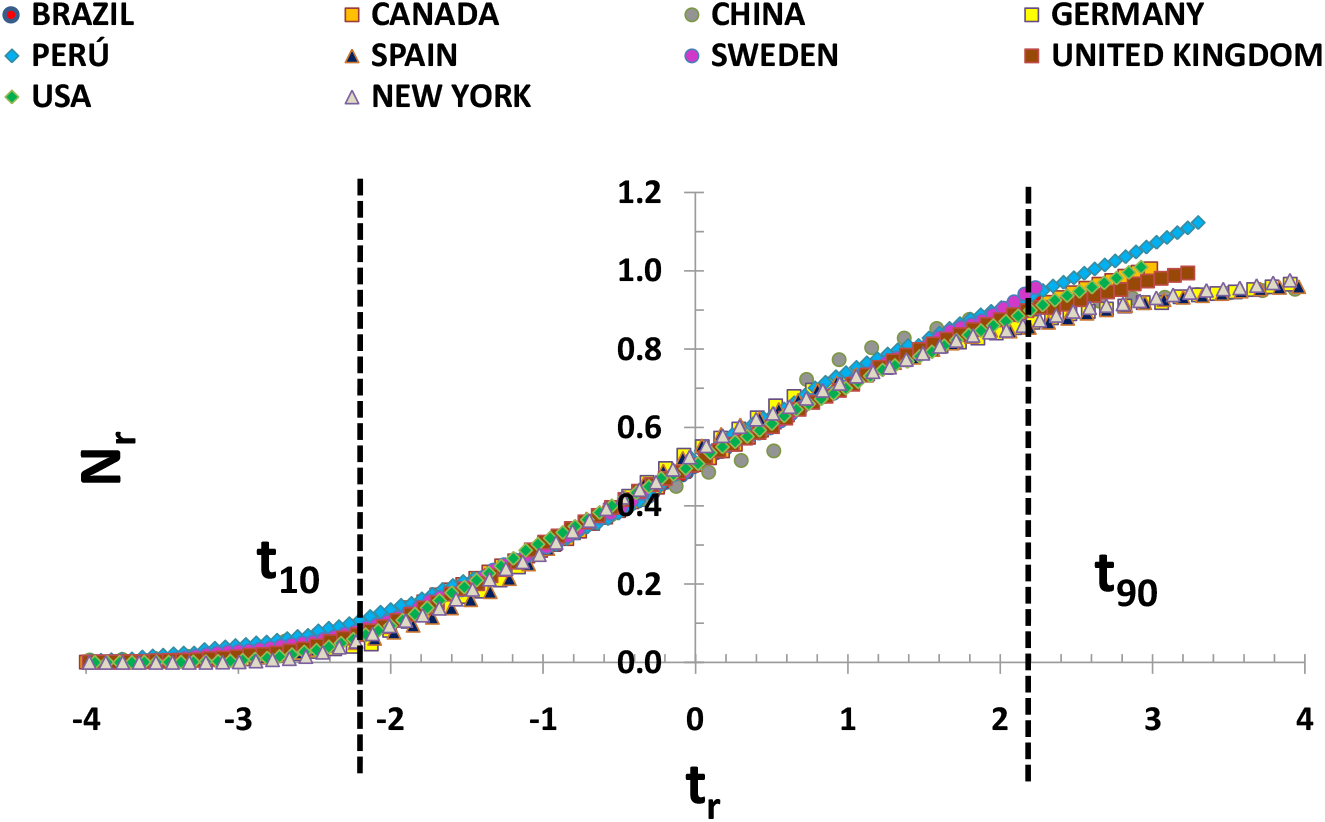
COVID-19 data shown in terms of the dimensionless variables *N*_*r*_ and *t*_*r*_, suggesting that the universal behavior of the F/D function appears to be present in the data from the selected countries. For reference, the *t*_10_ and *t*_90_ reduced times are indicated.

**FIG. 5:**
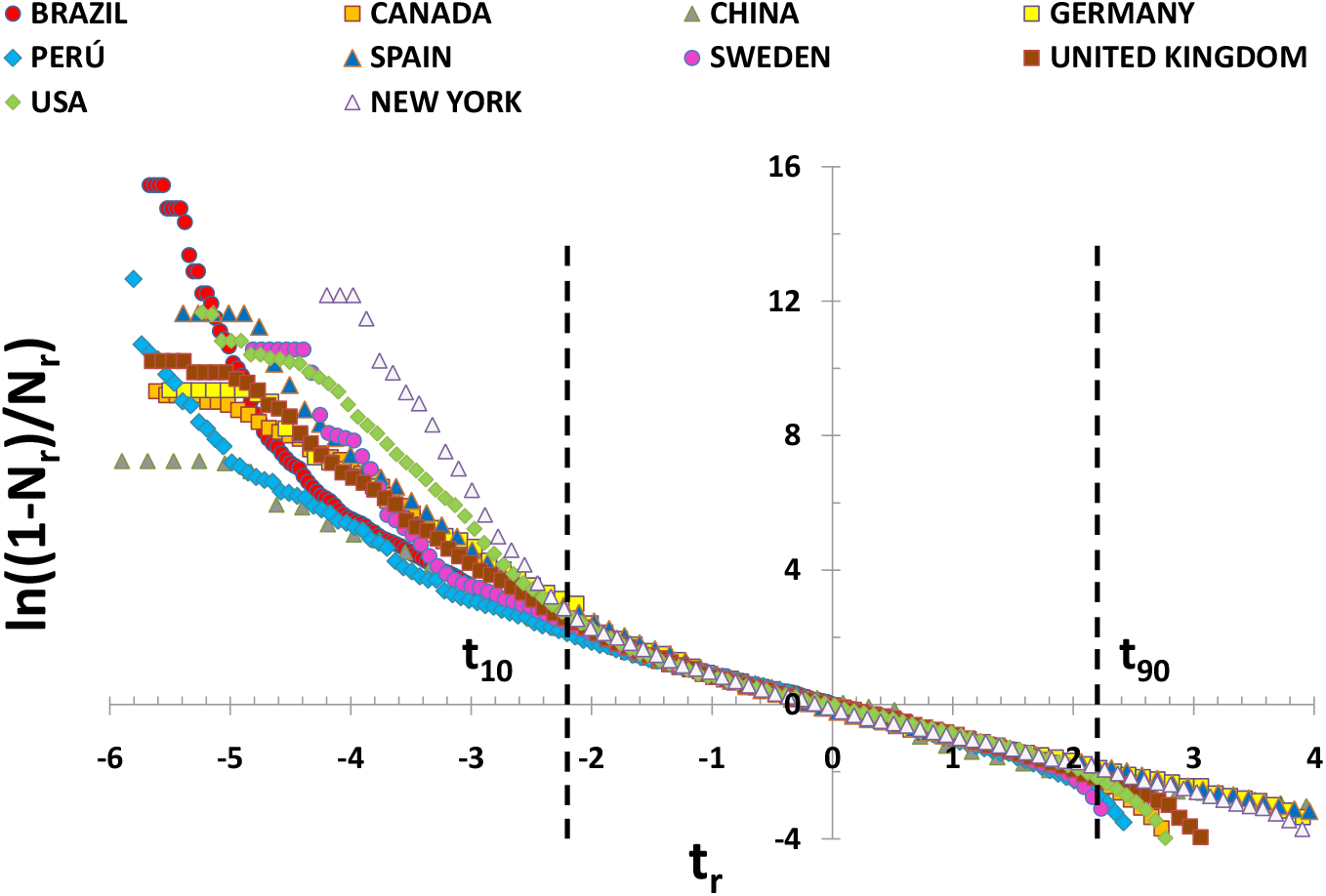
Similar to Fig. 4 but in the linearized version given in Eq. 6.

## IV. RESULTS AND DISCUSSION

Although the dynamics of the pandemic seems to have a universal behavior, it is clear from Figs. 2-5 that the data for the different countries do not strictly align on a single straight-line. There are many factors affecting the number of daily infected cases. Among them, the most relevant are those closely related to the measures adopted by the authorities of the different countries in an effort to control the disease, to reduce the number of cumulative infected cases, and to “flatten the curve”, keeping the *PR* below the available hospital resources at any given time to avoid overloading the system.

Nevertheless, by discussing a few examples, we will show how we can still use the linearized reduced forms, to correlate the data and shed light on the dynamics of the epidemic.

The first examples are devoted to China and Germany. The fits of Eq. 6 to the data are shown in Fig. 6 and give roughly slopes ≈ -1 and ≈ 0 intercept. For China, and for *t*_*r*_ ≥ 5 the slope is small. The value of *t*_*r*_ = 5 corresponds to *t*= 66 days (Mar-5) within the flattened part of the *N* (*t*) curve, meaning the epidemic was almost controlled. Among the countries selected in this study only China and Germany present this kind of behavior where the slope is ≈-1 and the intercept ≈ 0. We believe this is related to the fast measures taken by the chinese authorities to manage the spread of the disease (See Fig.1 in Ref. [13]). In the case of Germany the adopted strategy of massive testing, social distancing, and isolation of infected people since the early beginnings, was very effective to control the pandemic [14].

**FIG. 6:**
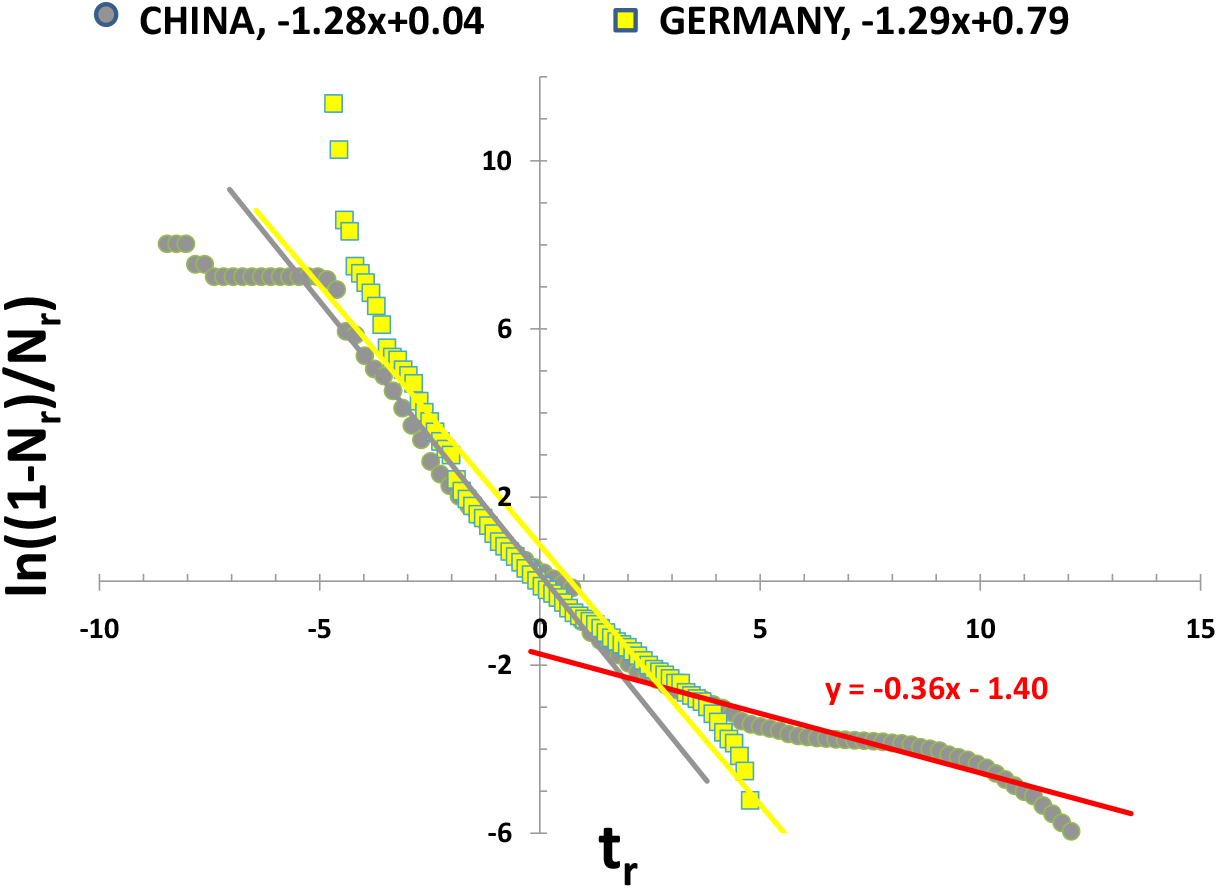
ln((1 − *N*_*r*_)*/N*_*r*_) is plotted vs *t*_*r*_ for the COVID-19 data of China and Germany. The equations for the straight line fits are given in the top legend.

Let us consider now Fig. 7, which presents in more detail the data in Fig. 5. Each country data set can be reasonably fit with two straight-lines, the universal region near *t*_*r*_ ∼ 0 and those corresponding to earlier reduced times. The slope of the straight lines is the inverse of the diffuseness Δ*T*, which measures the transmission rate *λ* in the SIS model. Therefore, a change of slope can be interpreted as a change in the transmission of the infectious disease. *A priori*, such a change appears contradictory to the universality conjecture given that the diffuseness is determined by fitting the F/D function, and therefore should be unique. However, the linearized forms are very instructive in the sense that it allows to investigate in more detail the development of the evolution of the epidemics and how external factors might influence it. In what follows, we will show that the universal behavior can be recovered. One may argue that presenting the data in this form, provides a more sensitive way to highlight potential deviations due to limited reporting, incomplete statistics, etc.

**FIG. 7:**
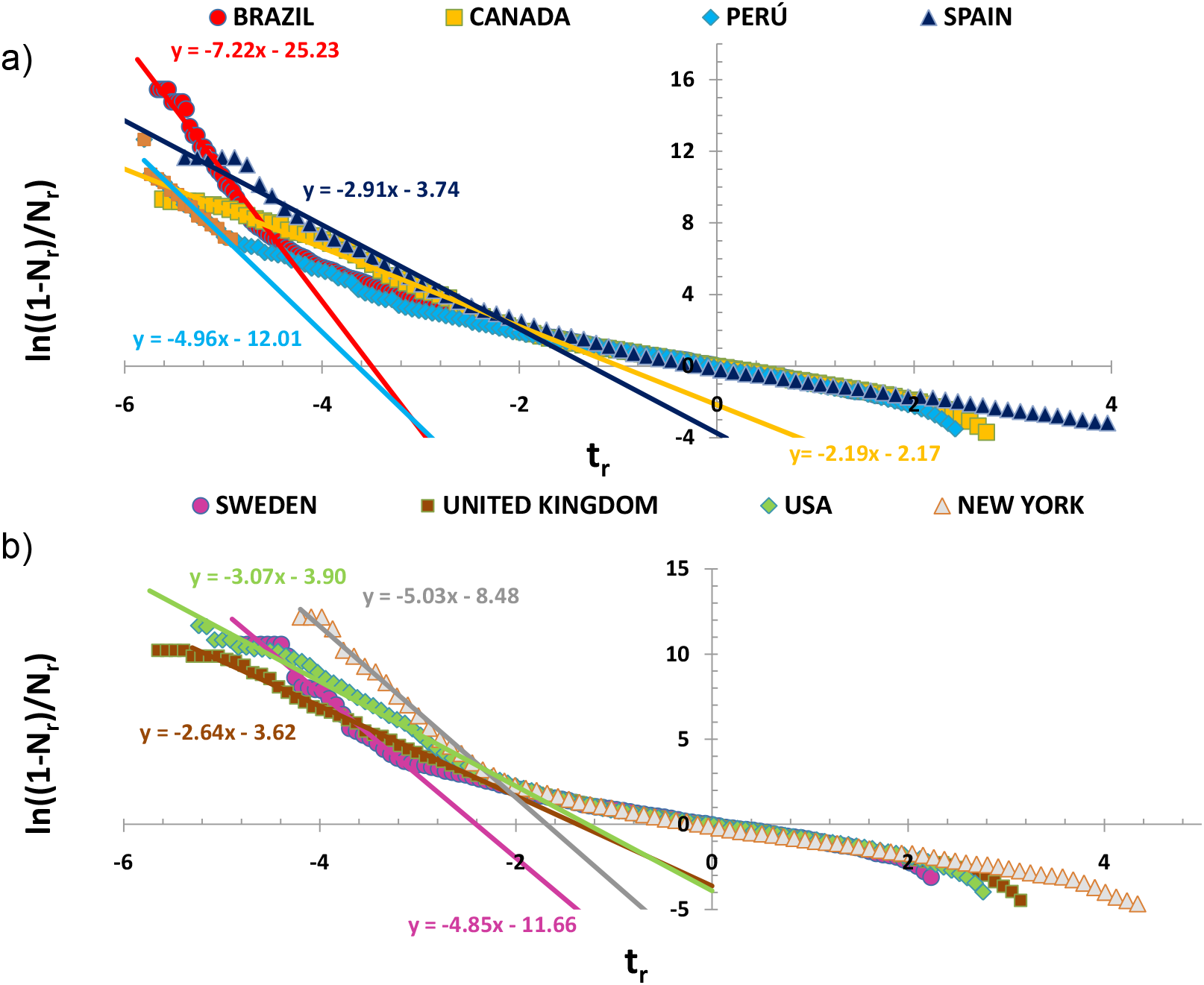
As in Fig. 6, for the data of the listed countries.

### A. *Prior* and *post* analyses

An inspection of Fig. 7 reveals also a linear dependence anticipated in Eq. 6 for times smaller than a characteristic time, *t*_*cross*_, the date at which the change of slope occurs for each country data set, just before the universal behavior sets in. The reduced slopes of the straight lines measure early transmission rates and suggest, for example, that in the case of Sweden this was ≈ 4.9 faster at the beginning of the pandemic, i.e. *t*_*r*_ *< t*_*cross*_. Similarly, in the USA that factor is ≈ 3.1, and so on for the rest of the countries. Changes in the slopes certainly reflect the way in which general living conditions are affected by the implementation of policies (travel restrictions, border closures, quarantines, social distancing, etc.) to control and limit the contagion. Consider the case of New York City, where the change of slope occurred on ≈ Mar-20. Social-distancing measures implemented on Mar-13 [15], must be reflected in a decrease in the number of daily infected cases and thus correlate well with the date of the sudden change in the slope.

Finally, let us come back to the discussion about the loss of universality in our approach. To that effect, we consider a slightly different analysis of the data. From Fig. 7 the date at which the sudden change of slope occurs, *t*_*cross*_, is determined for each country. Now, each set can be separated into two sub-sets: the first one referred to as *prior* starting on Feb-20 to *t*_*cross*_, and the second one, *post*, from *t*_*cross*_ to May-31, except for Brazil and Perú for which the analysis was extended to Sep-30 and Jul-20 respectively. Then, the same analytical F/D function is applied to each sub-set of data as presented in Figs. 8 and 9. The analyses of both *prior* and *post* data sets show that the average straight lines have reduced slopes: 1.15 ± 0.23 (*prior*) and 1.11 ± 0.08 (*post*), and intercepts: -0.21 ± 0.48 (*prior*) and 0.08 ± 0.09 (*post*) very close to the universal values.

**FIG. 8:**
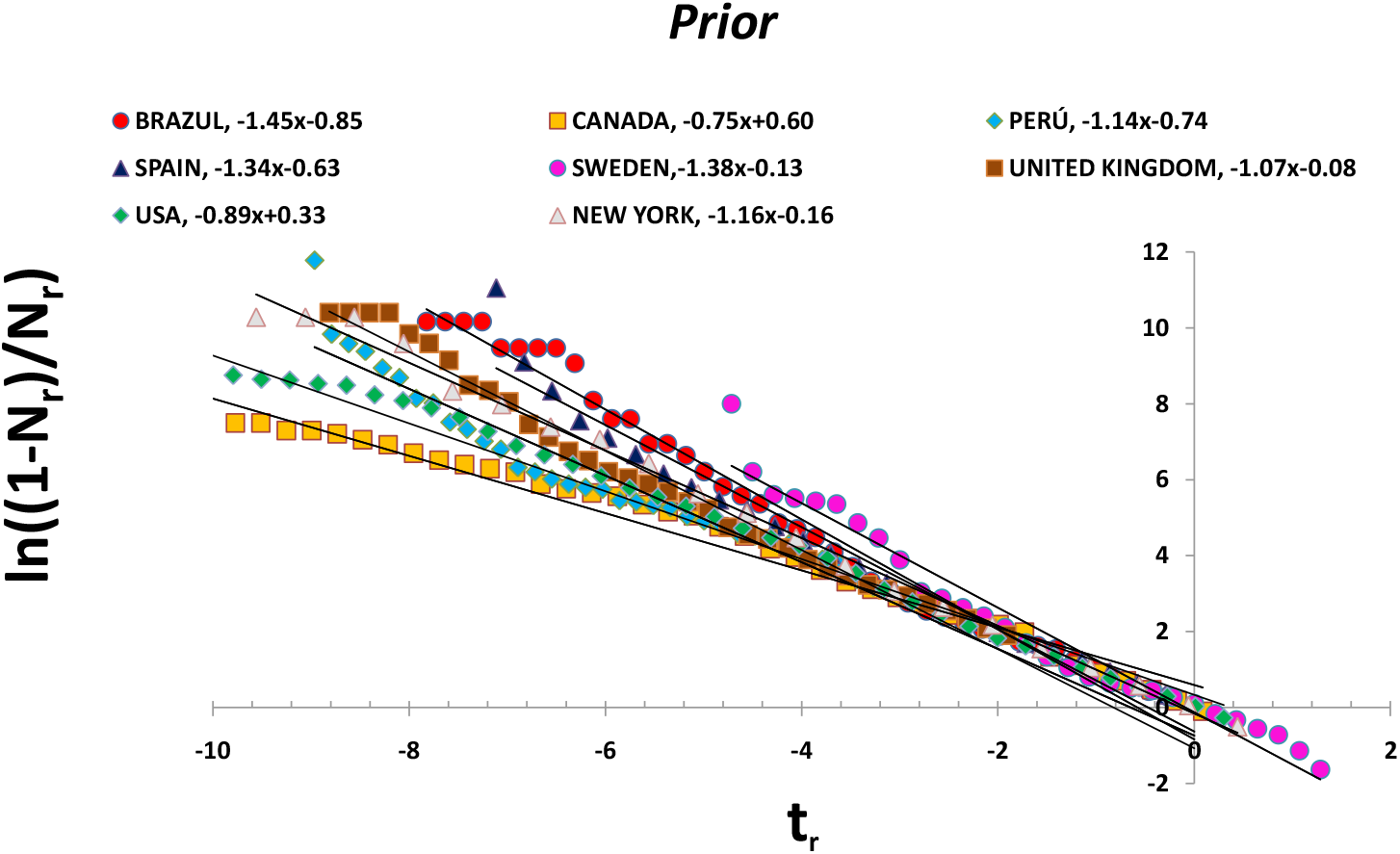
Plots of ln((1 − *N*_*r*_)*/Nr*) vs *t*_*r*_ for the *prior* sets of the selected countries. The linear fits are given in the legend.

**FIG. 9:**
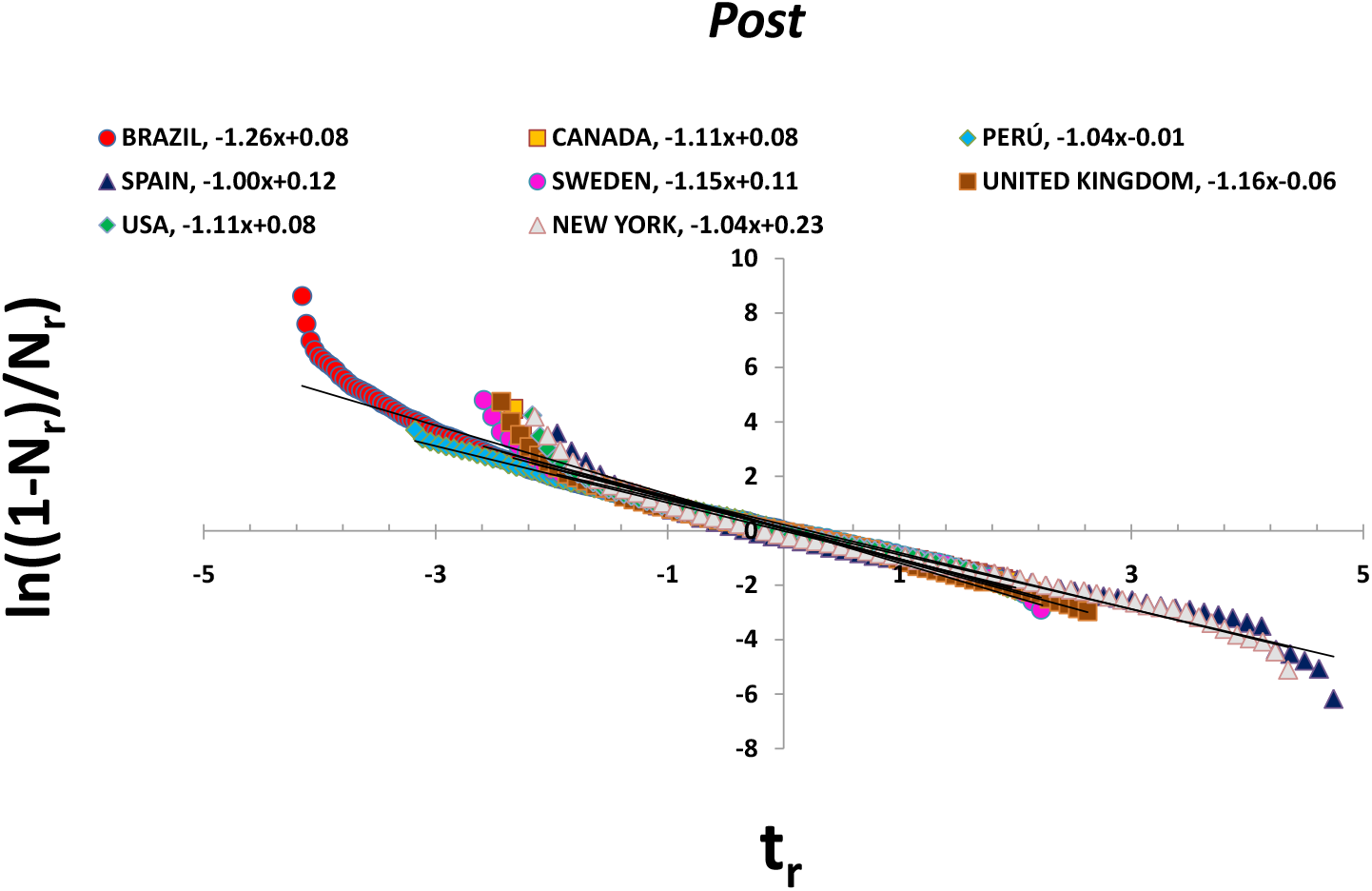
As in Fig. 8 but for the *post* data sets.

Thus, it seems that the empirical data are amenable of a description in terms of the linearized equations discussed above and follow a universal pattern in both *prior* and *post* analyses. It emerges from the comparison of these two sets that the implemented pandemic policies, aimed at reducing the impact of the COVID-19, have been able to decrease the basic reproduction number ℛ_0_ by a factor of ≈ 3, as measured by the ratio of the *prior* and *post* slopes 1*/*Δ*T*. Since ℛ_0_ ∼ 3, the reduction factor brings it down below the critical value of 1. This is shown, as an example, for the data reported in New York in Fig 10.

**FIG. 10:**
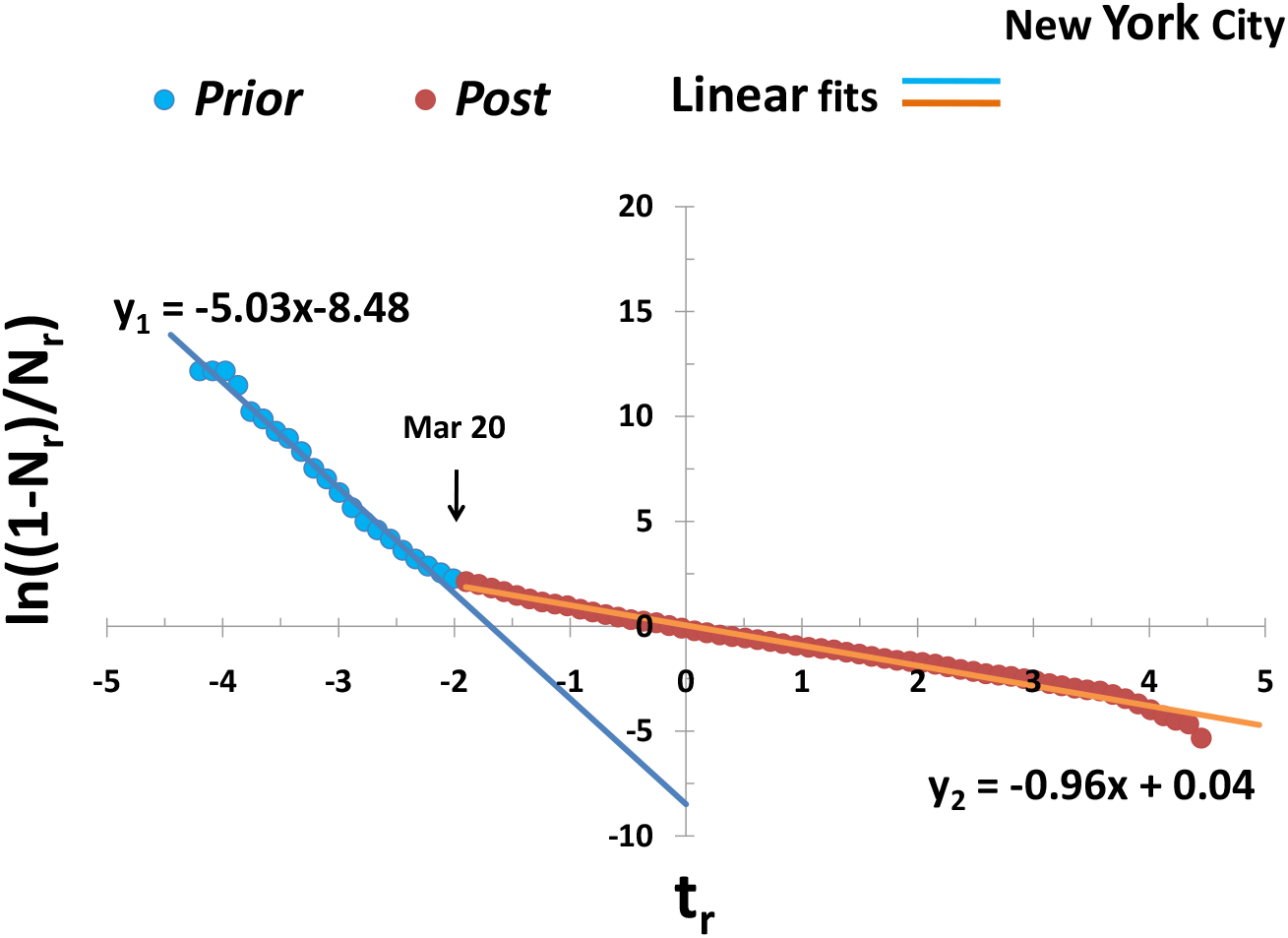
Plot of ln((1 − *N*_*r*_)*/Nr*) vs *t*_*r*_ for each subset of data *prior* and *post* reported in New York City.

### B. A benchmark with the extended SEICRD model

In this subsection we present the results of an extension of the SIR model to allow a time dependant infection rate, which in fact we model as a time dependant basic reproduction number ℛ_0_. Our objective here is to provide support of our simple phenomenological approach by comparing with the results of an established mathematical model, commonly used in epidemiological studies. In particular, we focus on the concept of the *t*_*cross*_ value and the *prior* and *post* analyses that follow.

In section I, we briefly presented the SIR model along with the system of differential equations governing the time evolution of the Susceptible, Infected and Recovered compartments. Ref. [16] presents a useful overview of this class of models, some of their theoretical properties and natural ways of extending them to more complex situations. Based on Refs. [16, 17] we thus modify the SIR model to account for individuals in the population that undergo different stages of the disease. We include an Exposed (E) compartment (those that have contracted the disease but cannot yet spread the virus), and two more compartments Critical (C) (those that need intensive care to model the overflow of Intensive Care Units (ICU) and Deaths(D) (cumulative deaths caused by the pandemic) and thus its name SEICRD. Further details are given in the Appendix.

In section IV A, the time *t*_*cross*_ was correlated to the implementation of policies that reduce ℛ_0_.We introduce in the SEICRD model a function of the form:

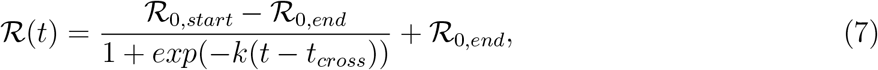

where the following desired properties are met:

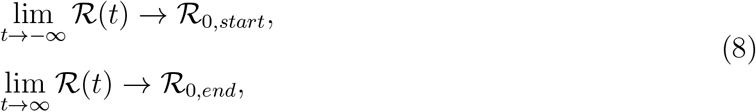

to simulate a change in the slope from ℛ_0,*start*_ to ℛ_0,*end*_ at *t* = *t*_*cross*_. The sharpness of the change is controlled by the parameter *k*. These additional parameters could either be fixed to suitable values, or fit to empirical data (See Appendix).

Here we just present the case of New York City. The SEICRD simulated data are contrasted with the F/D analysis in Fig. 11 that shows a plot of the dimensionless variables ln((1 - *N*_*r*_)*/N*_*r*_) vs. *t*_*r*_, to be compared with that of Fig. 10. The similar behavior of the universal straight lines and relevant dates is remarkable, suggesting that in spite of its simplicity, the approach captures the main ingredients of a more sophisticated model.

**FIG. 11:**
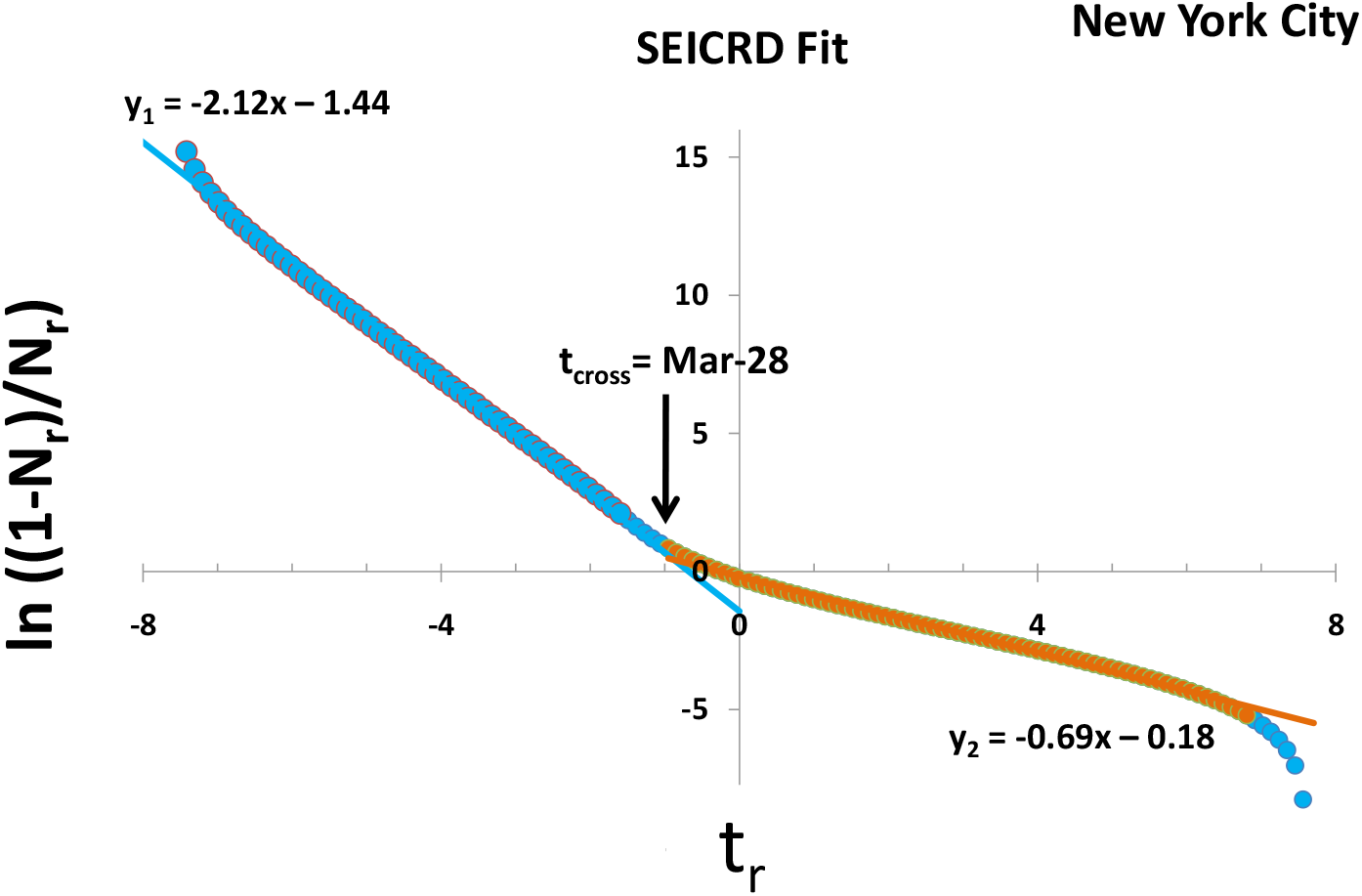
Plot of ln((1 − *N*_*r*_)*/Nr*) vs *t*_*r*_ for the simulated SEICRD data for New York City.

### C. Extended analysis beyond May-31

Due to the large extent of the pandemic, it is important to demonstrate if our analysis could also be applied to the second outbreak already seen in most countries and, specifically, if the concept of universality discussed in Section IV is still valid. The following example describes the analysis applied to the data reported in the United Kingdom.

An inspection of Fig. 12 a) allows us to identify two regions defined by a characteristic separation time *t*_*sep*_ between the peaks of the outbreaks. Each full data set can be then separated into two subs-sets: the first one, referred as to *Outbreak 1*, from Feb-20 to Jul-18 (*t*_*sep*_ for this case). The second subset, *Outbreak 2*, goes from Jul-19 until Nov-30. The same analytical F/D functions was then applied to each of the sub-sets giving the following parameters: 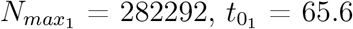, days (with respect to Feb-20) and Δ*T*_1_= 13.9 days for set 1 and 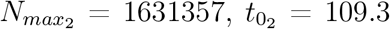, days (with respect to Jul-18) and Δ*T*_2_= 16.7 days for set 2. The results are summarized in Fig. 12 b) the reduced straight lines have slope and intercept close to -1 and 0 the universal values. The *prior* and *post* behavior discussed in Section IV A is also present in both outbreaks. Perhaps not surprising, the example confirms that the universal behavior is observed for the two outbreaks and anticipated to follow in subsequent ones. Of course, the same analysis can be easily applied to the extended data for other countries with similar results.

**FIG. 12:**
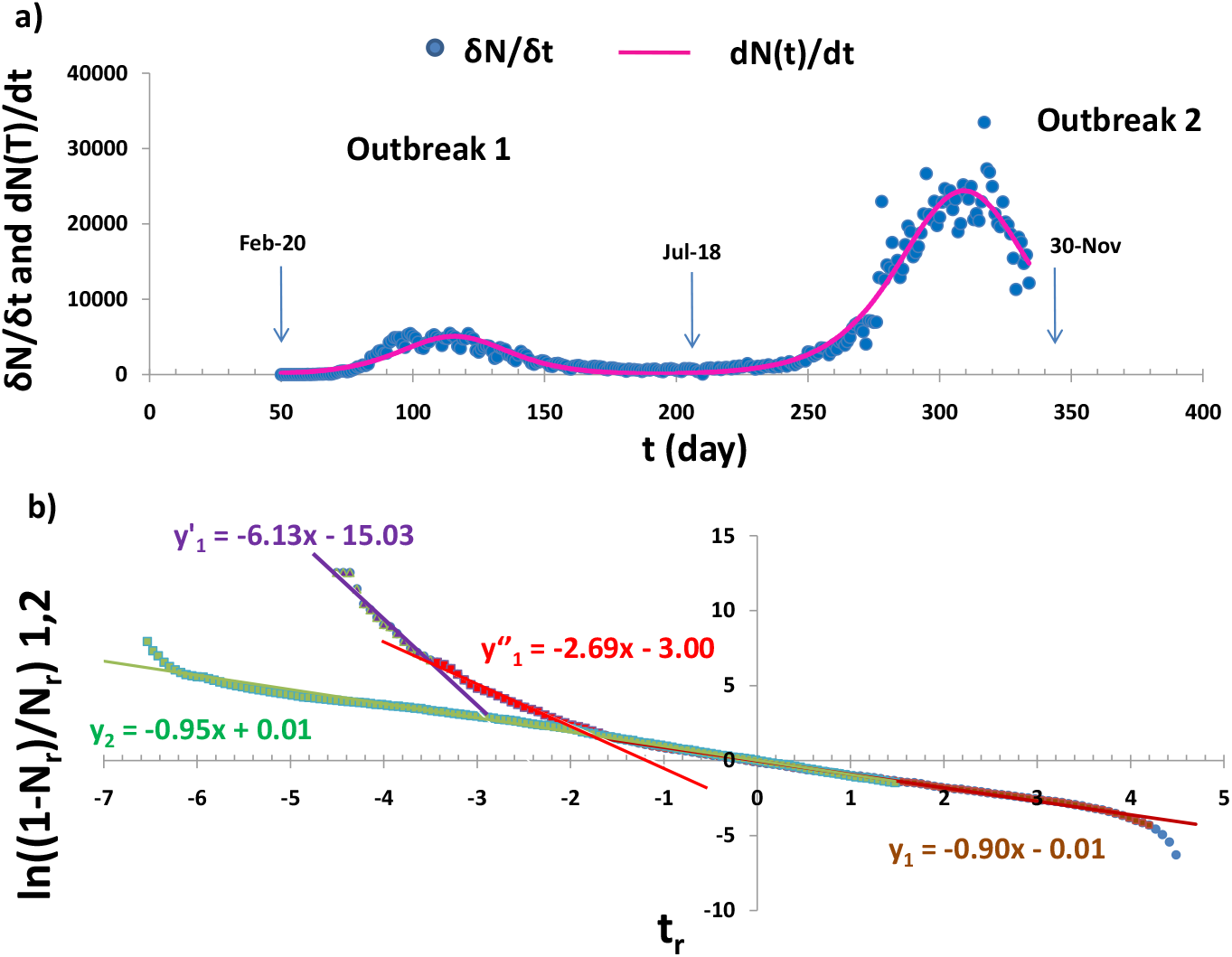
(a): Daily infection cases in the United Kingdom from Feb-20 until Nov-30. (b): Universal curves for the two outbreaks ending and starting on Jul-18 as indicated.

As a final comment we note that the proposed formalism can be applied instead to the number of COVID-19 related deaths as a function of time, which could be a more robust indicator of the impact of the pandemic.

## V. SUMMARY AND CONCLUSIONS

We have presented a phenomenological analysis of the time evolution of COVID-19 in some selected countries, using an analytical function of the Fermi/Dirac (Wood/Saxon) type. In spite of its simplicity, it appears that the proposed approach describes the empirical data relatively well. The extracted characteristic parameters allow, especially in the early stages of the epidemic, to make reasonable estimates readily comparable with the facts, and without resorting to more complex epidemiological models. Notwithstanding, the formalism is supported by the solution of the SIS model, an approximation of the SIR model and its extensions.

The linearized form of the F/D function suggests the introduction of dimensionless variables to correlate all the information in just one graph that reveals the universality of the phenomenon. Within our framework, we showed that the evolution of the epidemic can be easily tracked with changes in the values of the reduced slopes (*prior* and *post* analyses), reflecting measures and policies implemented to mitigate the spread of the disease. Although we focused our presentation on the first wave, we have shown that the phenomenology is also applicable to subsequent outbreaks. Finally, it did not escape us that it could be of interest to apply the formalism to the analysis of other viruses like influenza, which will be the topic of a follow-up work.

## Data Availability

Data available as indicated in the references.

## VI. ACKNOWLEDGEMENTS

This work is based upon work supported by the Director, Office of Science, Office of Nuclear Physics, of the U.S. Department of Energy under Contract No. DE-AC02-05CH11231 (LBNL). We would like to thank Dr. J. Fernández Niello for his comments on the manuscript.

## Appendix: The extended SEICRD Model

### 1. Introduction

In this appendix we discuss in more detail the extended SEICRD model which was used in Section IV B to benchmark the phenomenological analysis. This model is a natural extension of the SIR and the SEIR models [16]. The extended SEICRD model (in short SEICRD model) is obtained by the addition of two extra compartments: Critical (*C*) and Death (*D*). The first one accounts for individuals that need intensive care during the course of the disease and allows us to model the overflow of Intensive Care Units (ICUs) and the impact on the fatality rates. Individuals can only enter this compartment from an infected (*I*) state, from which they follow the cycle to *D* or *R*. The second compartment will hold the cumulative deaths caused by the pandemic, with the assumption that only individuals from *C* can die.

### 2. The differential equations of the model

The model dynamics is schematically shown in the flow diagram below, where the different compartments along with their state transitions are identified:

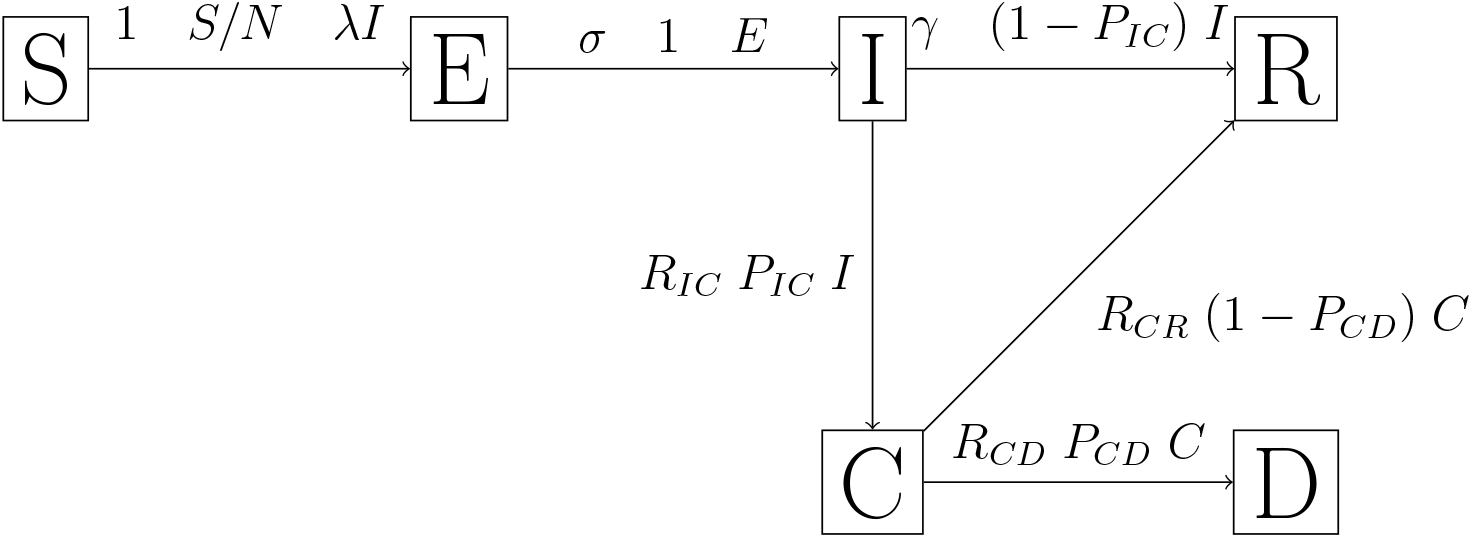

The system of differential equations readily follow:

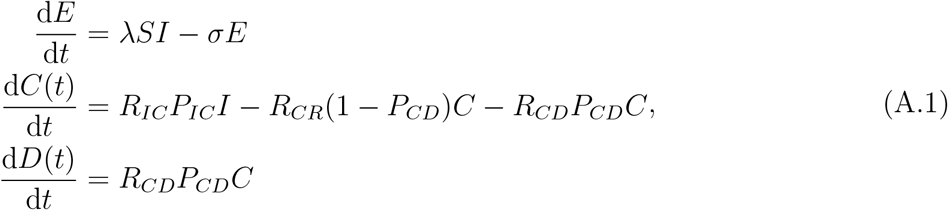

We further consider two additional and relevant features: a) the modeling of limited resources for ICUs; and b) the modeling of a time dependent infection rate.

The modeling of limited resources is done by a modification of the transition probabilities from *C* to *D* when the total number of patients in *C* is higher than the total number of ICUs available (*N*_*ICU*_). When such conditions are met, we assume the transition probability between *C* and *D* is for *C* - *N*_*ICU*_ individuals in *C*.

Finally, we introduce a time dependent infection rate into the model as a time dependent reproduction number ℛ(*t*). Our goal here is to give a robust model to support our interpretation of the *t*_*cross*_ value introduced in sub-sections IV A, IV B). Thus, we propose that ℛ(*t*) is given by a function of the form:

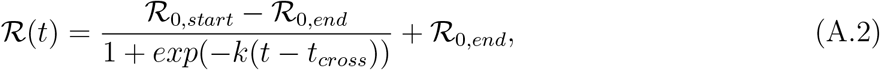

where the following desired properties are met:

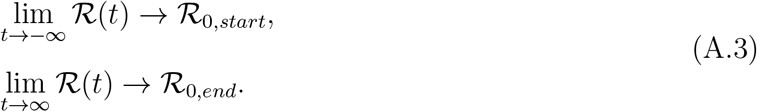

The sharpness of the change in slope, from ℛ_0,*start*_ to ℛ_0,*end*_ at *t* = *t*_*cross*_, is controlled by the parameter *k*.

### 3. The solution of the model equations

Following from the discussions above, the final system of differential equations is:

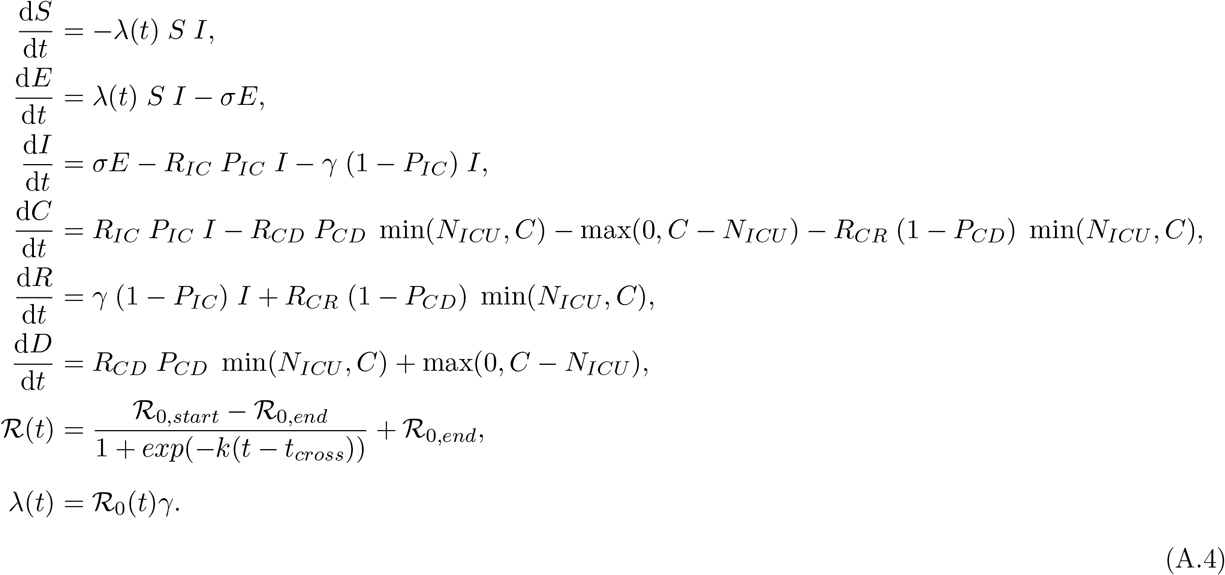

At first glance, we have several parameters we have to deal with. Nonetheless, some of them that do not have a significant variation during the process of the pandemic can be fixed, while the rest have to be fitted to the available data. In the following list the whole set of parameters and how they are treated in the fitting procedure are given:

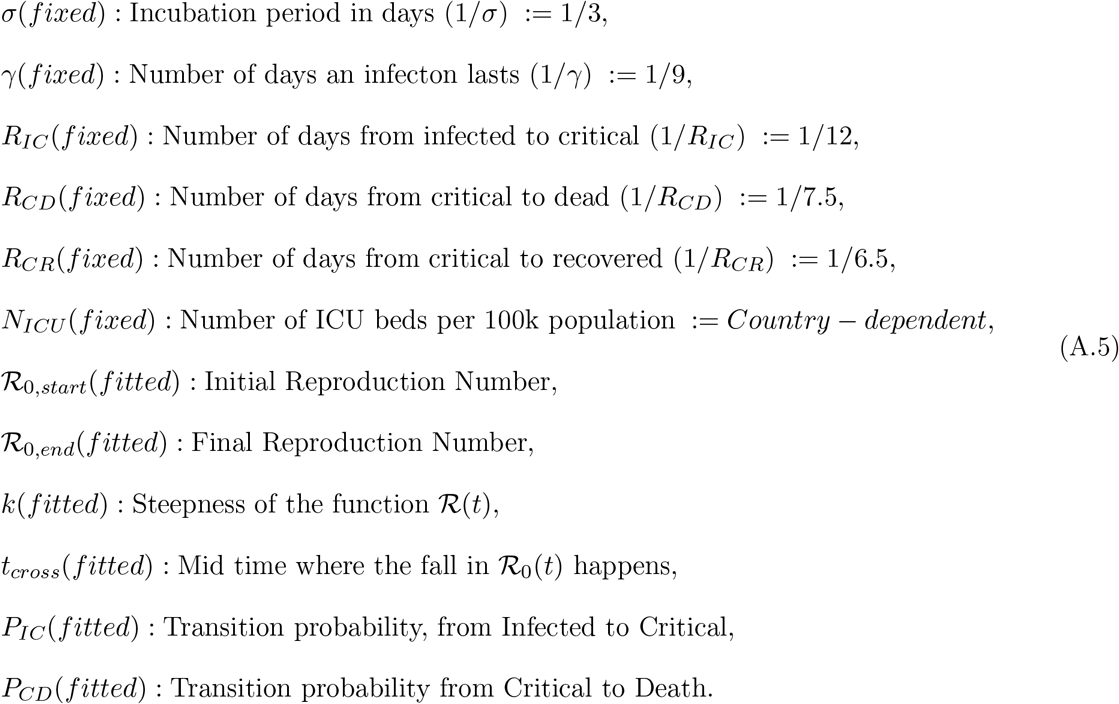

The values for *N*_*ICU*_ are not pandemic but country specific and were compiled from Refs. [18, 19] to get the most up-dated values.

A Python-code [20] was developed to solve equations (A.4) and fit the results to the available cumulative deaths data reported for the different countries. While fitting the model to the available reported deaths, the obtained evolution of the infected compartment is likely to reflect the total numbers of cases more precisely than if derived from the positive tested ones. The fit results are summarized in Table II.

**TABLE II:**
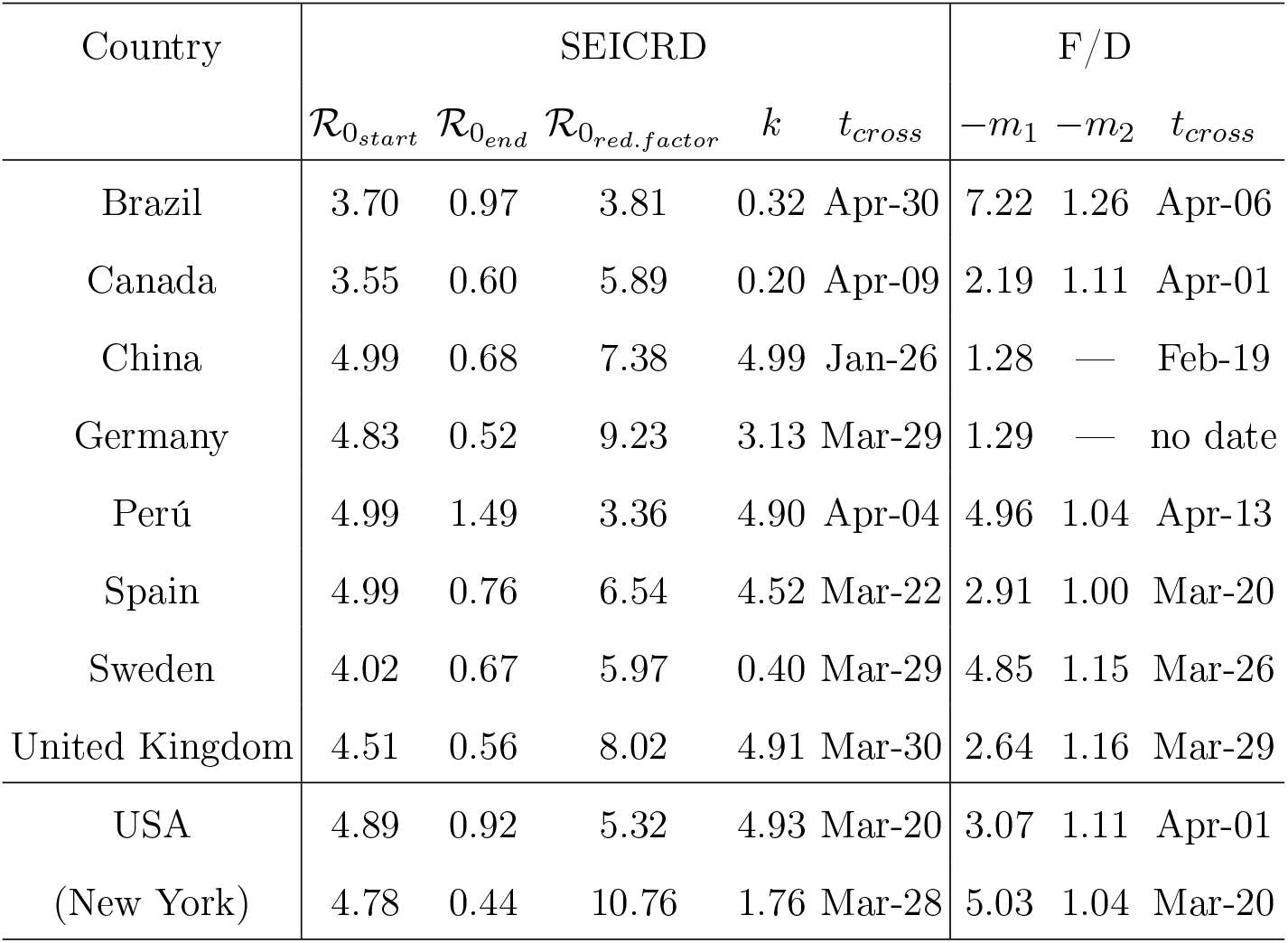
Results of the SEICRD model fit to the data for the countries considered in this work. The cut-off date for these calculations was set on Oct-29. The *m*_1_ and *m*_2_ values are extracted from Fig. 7 and Fig. 9 respectively, those for China and Germany from Fig. 6.

We have discussed in Section IV A that the linearized form of Eq. 6, fitted to the SEICRD results for New York City, captures the main ingredients of the evolution of the pandemic as predicted by a more realistic, yet more complex, model. A similar conclusion can be drawn from the analysis of the other countries.

## Footnotes

1 Numbers as of Dec-10.

2 All data used in this work have been extracted from the online reference sites [3, 4].

3 The number of coronavirus cases in the city of Wuhan, where the pathogen was first detected, may have been 10 times higher than official figures suggest, according to a recent Chinese CDC report.

